# The scale and dynamics of COVID-19 epidemics across Europe

**DOI:** 10.1101/2020.06.26.20131144

**Authors:** Christopher Dye, Russell C.H. Cheng, John S. Dagpunar, Brian G. Williams

## Abstract

The number of COVID-19 deaths reported from European countries has varied more than 100-fold. In terms of coronavirus transmission, the relatively low death rates in some countries could be due to low intrinsic (e.g. low population density) or imposed contact rates (e.g. non-pharmaceutical interventions) among individuals, or because fewer people were exposed or susceptible to infection (e.g. smaller populations). Here we develop a flexible empirical model (skew-logistic) to distinguish among these possibilities. We find that countries reporting fewer deaths did not generally have intrinsically lower rates of transmission and epidemic growth, and flatter epidemic curves. Rather, countries with fewer deaths locked down earlier, had shorter epidemics that peaked sooner, and smaller populations. Consequently, as lockdowns are eased we expect, and are starting to see, a resurgence of COVID-19 across Europe.

**One Sentence Summary:** A flexible empirical model shows that European countries reporting fewer COVID-19 deaths locked down earlier, had shorter epidemics that peaked sooner, and smaller populations.

## Main Text

The total number of COVID-19 deaths reported by European countries up to 31 July varied more than 100-fold, from approximately 100 in Croatia to more than 45,000 in the United Kingdom. In terms of the dynamics of coronavirus transmission, there are broadly three possible reasons why a country might suffer relatively few deaths. The first is that the transmission rate of the coronavirus, SARS CoV-2, is intrinsically lower in some countries, for example because infectious and susceptible individuals come into contact less frequently in less dense populations. This would be reflected in a relatively low value of the basic case reproduction number, *R_0_*. Figure 1a shows how *R_0_* changes the shape and scale of an epidemic, aided by a dynamic SEIR epidemiological model (Supplementary Materials), and with reference to deaths reported from Germany. Given a basic case reproduction number of *R_0_* = 3, the number of deaths reached a maximum of approximately 1600 in the week of 16 March, and an estimated total of 9300 people died from COVID-19 by the end of July (Figure 1a). If *R_0_* had been greater at the outset (*R_0_* = 6) the epidemic would have been larger (9800 deaths) and shorter, growing faster and peaking sooner. If *R_0_* had been lower (*R_0_* = 2), the epidemic would have been smaller (7900 deaths) and longer, growing more slowly with a delayed peak. A lower value of *R_0_* mitigates transmission and flattens the epidemic curve, protecting both health and health services (*1-7*).

**Fig. 1.**
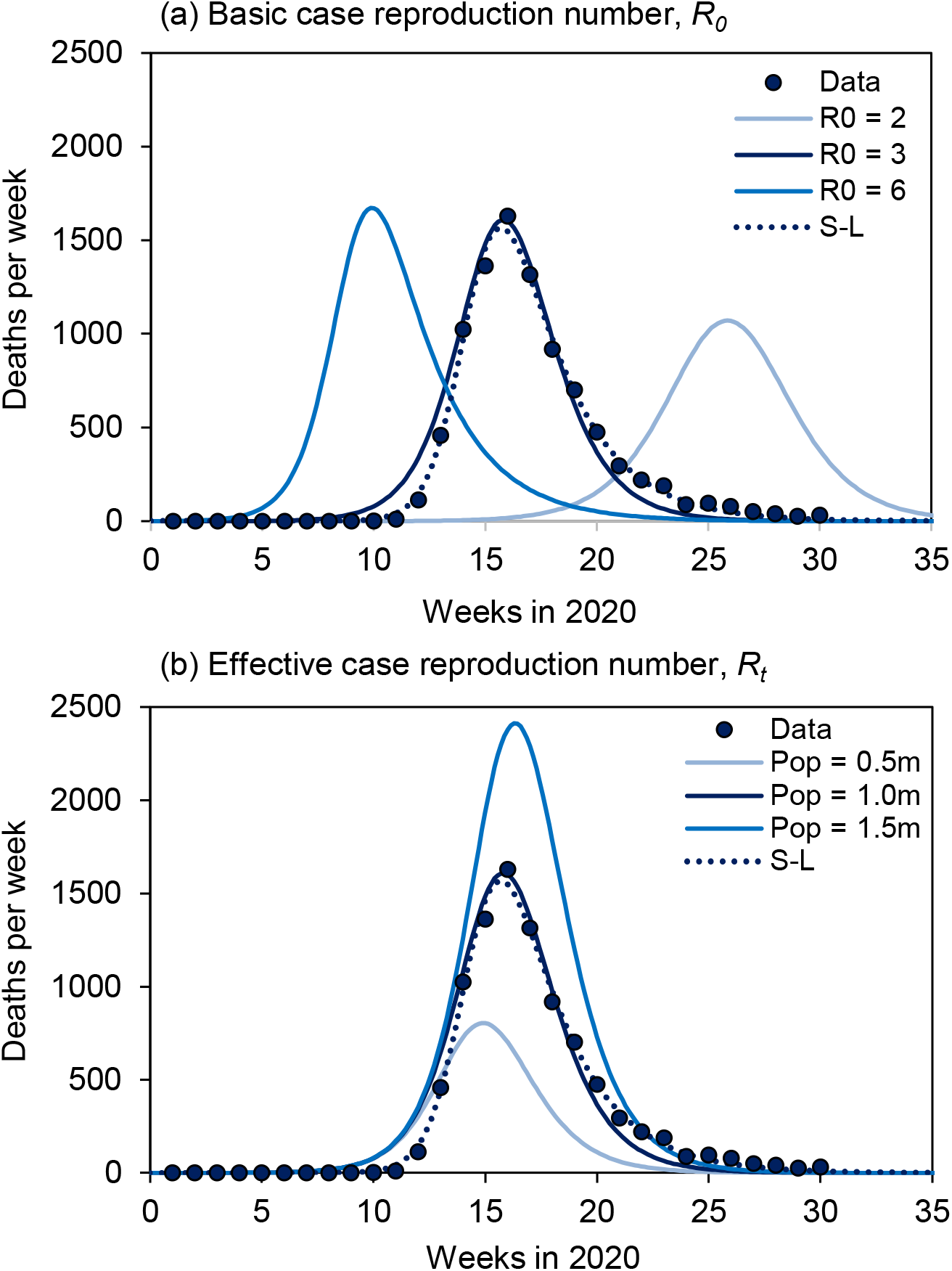
A SEIR transmission model (solid lines) illustrates the mechanisms that determine the scale and dynamics of COVID-19 epidemics, here described by the number of deaths reported each week. (a) Lower values of the time-invariant, basic case reproduction number (*R_0_* = 6, 3 or 2, adjusted e.g. by social mixing) mitigate transmission and flatten the epidemic curve. With lower *R_0_*, the maximum number of deaths per week is reduced and delayed, generating epidemics that are smaller and longer. (b) After the initial period of epidemic growth (governed by *R_0_*), the decline of the effective case reproduction number, *R_t_* < *R_0_*, is accelerated by limiting the size of the susceptible population (1.5, 1.0 or 0.5 million people), generating epidemics that are smaller and shorter, peaking earlier. The epidemics in (a) or (b) could also be replicated by reducing *R_0_* to R*_0c_* before or during the epidemic e.g. through non-pharmaceutical interventions. Panels (a) and (b) also show weekly reported deaths in Germany (points) described by the SEIR model and, for comparison, the skew-logistic model (dotted line; see Figure 2). The SEIR model and data (*38*) are described further in the Supplementary Materials.

The second possibility is that some countries have suffered fewer deaths because, after the initial period of maximum epidemic growth (governed by *R_0_*), the effective case reproduction number (*R_t_* ≤ *R_0_*) declined relatively quickly through time in a smaller populations of susceptibles, depleted by the build-up of herd immunity (*8*) (Figure 1b). This is a simple representation of heterogeneous exposure or susceptibility to infection which, compared with homogeneous exposure or susceptibility, would lower the fractions of people who become infected, ill and die (*9, 10*). Countries with fewer people exposed or susceptible to infection are expected to have smaller and shorter COVID-19 epidemics (Figure 1b).

The third possible reason for fewer deaths is that the basic reproduction number, *R_0_*, was reduced to a lower value, *R_0c_*, by control methods imposed before or during the epidemic. These are predominantly “non-pharmaceutical interventions” (NPI), which include personal (physical distancing, isolation, quarantine, hand hygiene, face covering), environmental (surface cleaning and ventilation), and social (travel restrictions, school and workplace closures, restriction on mass gatherings) methods of preventing contact and transmission between infectious and susceptible individuals. In this model of epidemic control, the case reproduction number is reduced through time, not because susceptibles are progressively depleted in an exposed subpopulation, but because they can no longer be contacted by infectious cases (*3, 11, 12*). A change from *R_0_* to *R_0c_* could replicate the effects seen in either Figure 1a or 1b, dependent on the timing and magnitude of the change, but independent of population size (Figure 1b). A consequence of this model of epidemic control is that, if NPI are relaxed in a population where many people are still susceptible to infection, we expect a resurgence of COVID-19.

Which of these three possibilities explains the variation in COVID-19 deaths among European countries? In this study, rather than presuming which mechanism of epidemic control is more likely *a priori*, we began by developing a flexible, empirical model (skew-logistic) to characterize the scale and dynamics of COVID-19 epidemics in European countries (Supplementary Materials). Unconstrained by biological assumptions, and agnostic to the mechanisms of epidemic control (Figure 1), the skew-logistic model allowed us to measure, independently of each other, the key components of an epidemic: the rate of epidemic growth, the size of the epidemic peak (maximum number of deaths per week), the period or duration of growth (estimated time from one death to the maximum number of weekly deaths), and the rate of decline. We then asked which of these elements best explains the differences among European countries, and by what mechanism (Figure 1).

The skew-logistic model gives an excellent description of COVID-19 epidemics in 24 European countries, which is generally better than the SEIR model without adjusting initial conditions (Figures 1 and 2, Supplementary Materials). The average initial growth rate of epidemics was 0.28/day (95%CI 0.006/day), doubling time 2.5 days), the average period of epidemic growth was 37 days (95%CI 4.0 days), and the average rate of decline was -0.05/day (95%CI ± 0.001/day, halving time 14.1 days). For each of the 24 countries, maximum likelihood estimates for parameters of the skew-logistic are tabulated in the Supplementary Materials.

**Fig. 2.**
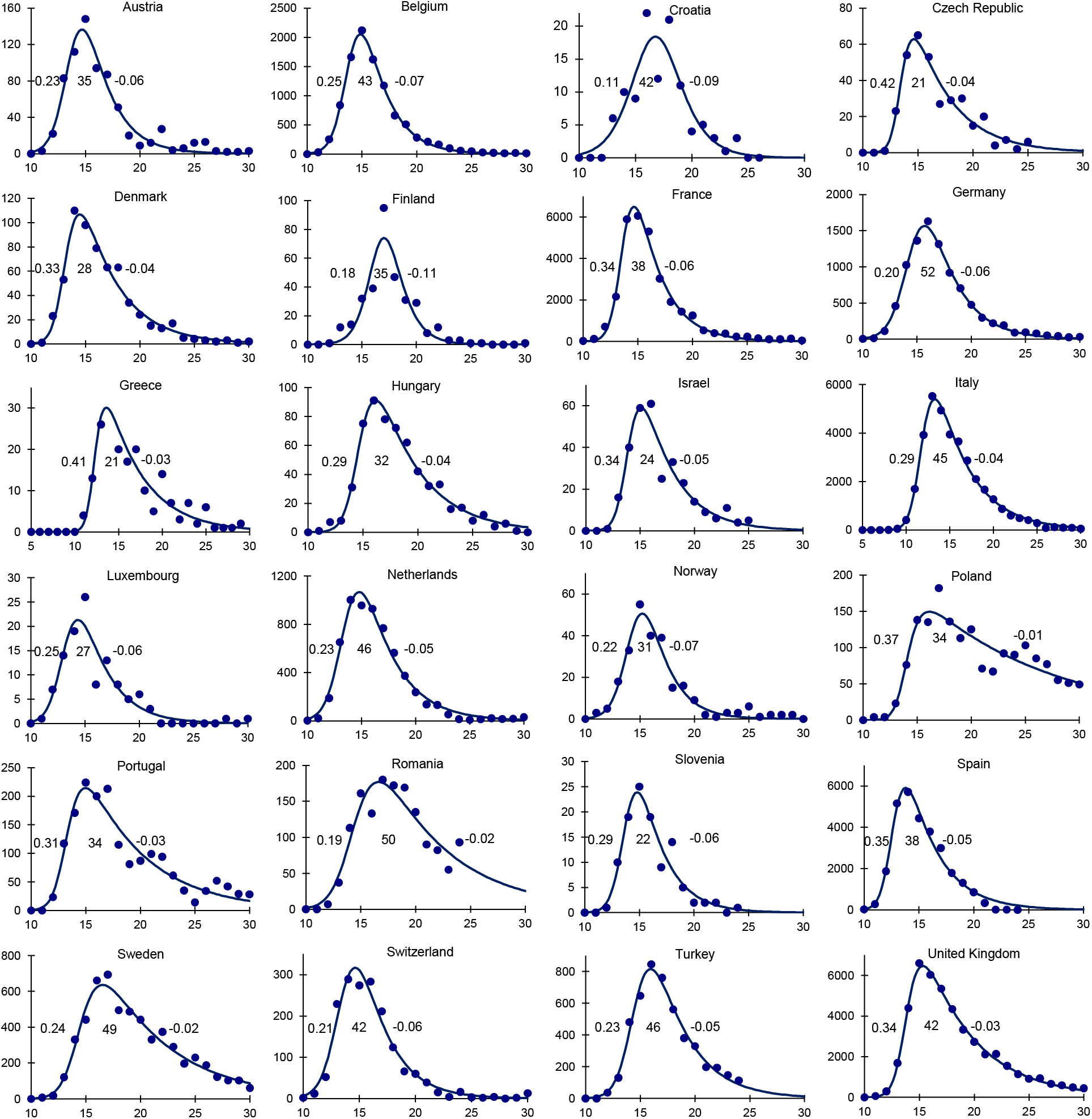
Epidemic curves for 24 European countries based on the number of deaths (points, vertical axis) reported each week (horizontal axis), described by the skew-logistic model (line) which is used to calculate the epidemic growth rate, the period of epidemic growth and the rate of decline in each country (inset figures, left to right). Although deaths are reported daily from each country, and growth rates and periods are given in days, the model is fitted to the weekly numbers of deaths in order to sum over the typical 7-day reporting cycle (Supplementary Materials). Data source (*38*).

The skew-logistic model shows how the number of COVID-19 deaths was associated with these 10 characteristics of the epidemics and the size of national populations. In general, countries reporting fewer COVID-19 deaths had fewer inhabitants (*t* = 6.83, *p* < 0.001; Figure 3a). In countries reporting fewer deaths, epidemics neither grew more slowly (Figure 3b) nor declined more quickly (Figure 3c). Rather, epidemics that caused fewer deaths were curtailed; they grew for shorter periods of time (*t* = 3.91, *p* < 0.001; Figure 3d). And shorter epidemics were smaller 15 epidemics, even though shorter epidemics tended to increase more quickly (*t* = 3.95, *p* < 0.001; Figure 3e).

**Fig. 3.**
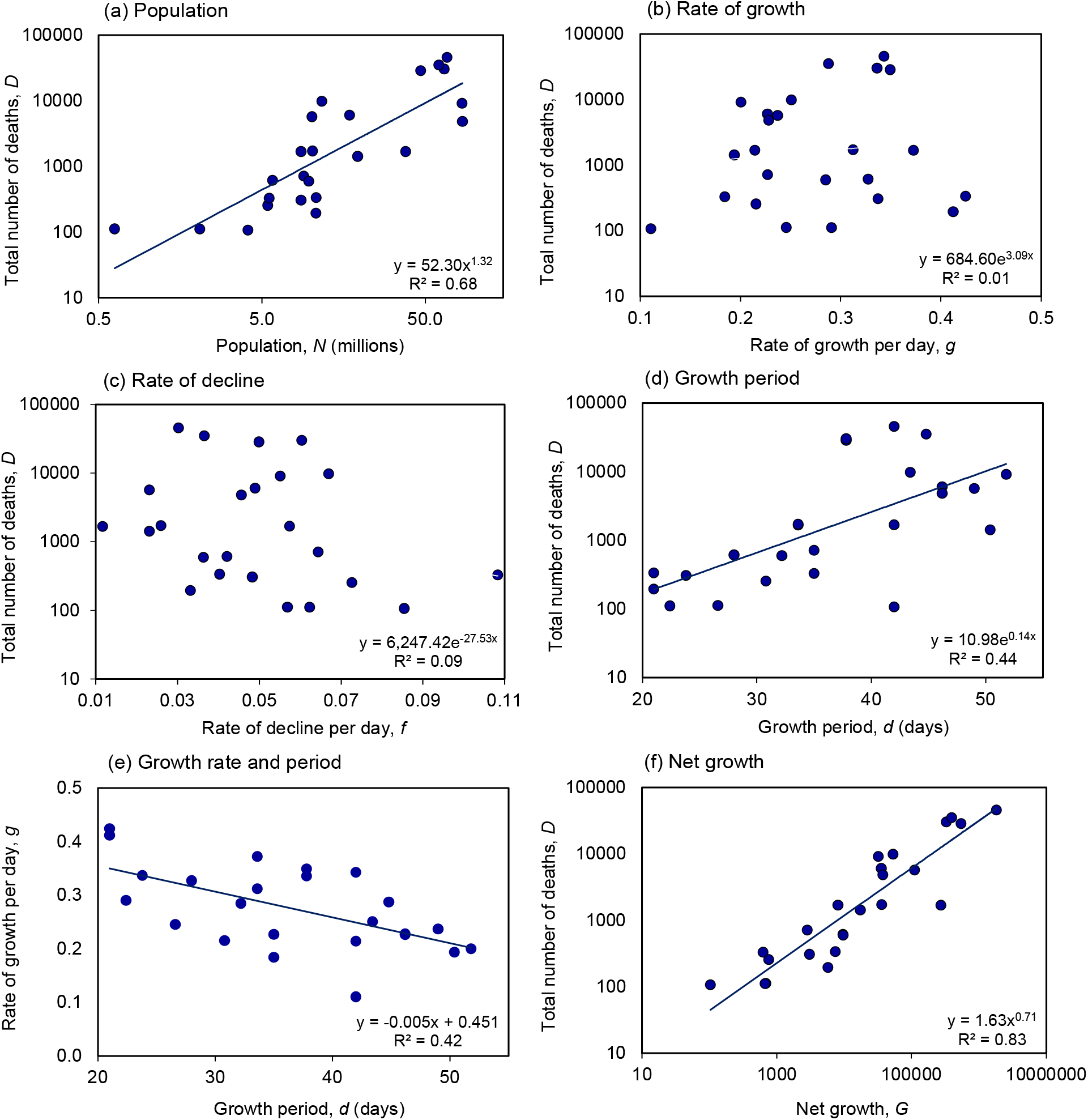
Determinants of the total number of reported COVID-19 deaths in each of 24 European countries, using estimates derived from the skew-logistic model (Figure 2). (a) The number of deaths increased with national population size. The number of deaths varied independently of (b) the rate of epidemic growth and (c) decline. But (d) more deaths were reported when epidemics were allowed to grow for longer, even though (e) longer periods of growth were associated with lower epidemic growth rates. (f) The number of deaths was strongly associated with growth rate and period combined as net growth, *G = e^gd^*. Each panel gives the regression equation (regression line shown if statistically significant) and the fraction of the variation explained by each independent variable (*R^2^*). Data source (*38*).

The product of the rate (*g*) and period of growth (*d*) defines net epidemic growth — how much an outbreak expands in size during the growth period. Net growth, measured by *G = e^gd^*, accounted 20 for 83% of the inter-country variation in the number of reported deaths (*t* = 12.4, *p* < 0.001; Figure 3f). Although such a relationship is expected in principle, it is surprising that theory is upheld so faithfully in data collected in different ways across 24 diverse European countries.

Drawing together the data across these 24 countries (Figure 3), multiple regression analysis 25 shows that both population size (*N*) and net growth (*G*) were strongly and independently associated with the total number of deaths (*D*), such that ln (*D*) *=* 0.31 ln (*N*) *+* 0.67 ln (*G*). The number of deaths therefore increased logarithmically (less than proportionally) with population size (*D ∝ N*^0.31^, 0.31 < 1, *t* = 4.1, *p* < 0.001) and net growth (*D ∝ G*^0.67^; 0.67 > 0, *t* = 13.9, *p* < 0.001; overall *R^2^* = 0.83) and, given the definition of G, exponentially with epidemic growth rate 30 and period (*D ∝ e*^0.67^*^gd^*). The finding that *D ∝ N*^0.31^ indicates that the number of inhabitants of a country is not simply an epidemic scaling (proportionality) factor with effects that can be captured as deaths per capita, as implied in some presentations of the data (*13, 14*).

Among competing explanations for the large inter-country variation in reported deaths, these 35 data are consistent with the view that there were systematic national differences in the effective case reproduction number (*R_t_* and/or *R_0c_*, Figure 1b) but not the basic case reproduction number (*R_0_*, Figure 1a). What, then, were the relative roles of *R_t_* and *R_0c_* as determinants of the numbers of deaths?

Because the period of epidemic growth (Figure 3d) and net growth (Figure 3f) accounted for much of the inter-country variation in reported deaths, the timing of interventions is expected to influence the total number of deaths, moderated by population size (Figure 3a). One summary measure of non-pharmaceutical interventions is the COVID-19 Containment and Health Index (CHI), a composite metric based on eleven indicators including containment and closure policies 45 (schools, workplaces, travel bans) and health system policies (information, testing, contact tracing), with a score for each country varying between 0 and 100% (Supplementary Materials) (*15*).

Figure 4a shows that the total number of deaths (*D*) reported from 30 countries (the 24 in Figures 2 and 3, plus Albania, Bulgaria, Iceland, Ireland, Serbia and Slovakia) was closely associated with the number predicted on the basis of population size (*N*) and the cumulative number of deaths that had been reported by the date of lockdown (*D_L_*), as judged from the CHI (different interpretations of the time of lockdown give similar results; Supplementary Materials). In Figure 4a (similarly to Figure 3f), the number of deaths increased logarithmically with population size (*D ∝ N*^0.49^, 0.49 < 1, *t* = 5.3, p < 0.001) and with the number of deaths at lockdown (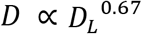; 0.67 > 0, *t* = 3.4, p < 0.002; overall *R^2^* = 0.84). Thus, to a good approximation, the number of deaths in each country varied in proportion to *N*^1/2^ and 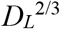.

**Fig. 4.**
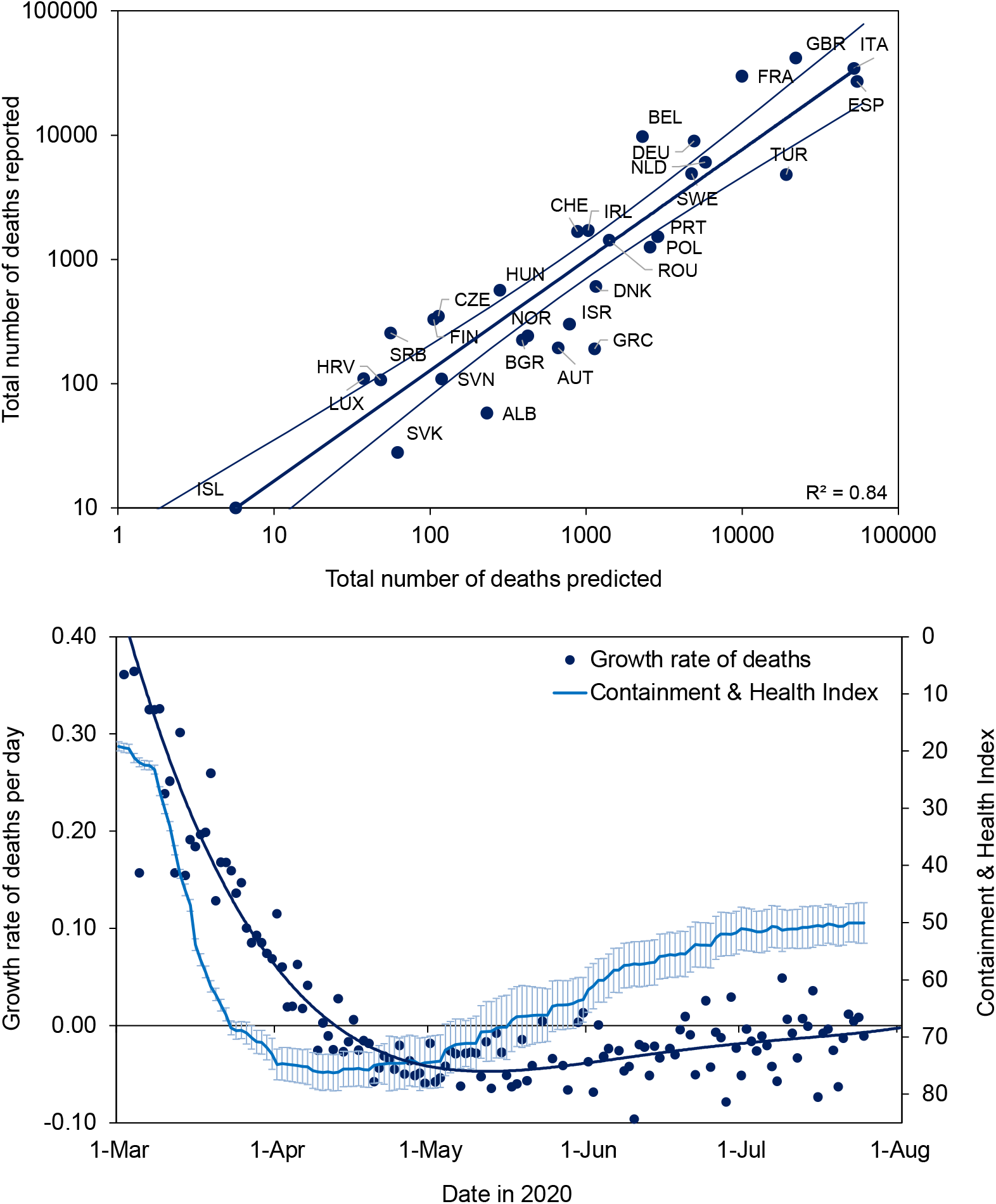
(a) Total numbers of COVID-19 deaths (*D*) reported from 30 European countries (the 24 in Figures 2 and 3 plus Albania, Bulgaria, Iceland, Ireland, Serbia and Slovakia) in relation to the number of deaths predicted from population size (*N*) and the number reported at the time of lockdown (*DL*). Errors on the regression line mark 95%CI. (b) The imposition and release of lockdown across 30 countries, as measured by the mean value of the Containment and Health Index (light blue, error bars 95% CI, reverse scale on *y*-axis), which was followed by the changing daily growth rate of deaths (dark blue points, fitted polynomial line of order 4). Further information and analysis, including full country names, are in the Supplementary Materials. Data sources (*15, 38*).

Considering all 30 countries together, lockdown evidently stopped and reversed the increase in COVID-19 deaths in European countries (Figure 4b). Most of the 30 countries investigated here locked down within a short period in March and the CHI surpassed 70% on average by 16 March. Lockdown was followed by a progressive fall in the daily growth rate of deaths (daily change in the 7-day running mean), so that deaths were no longer increasing (negative growth) by the second week in April.

Just as the lockdown appears to have prevented the spread of COVID-19, the easing of restrictions preceded a resurgence. The average value of the CHI peaked at 75% on 12 April and was held above 70% for 46 days. As lockdowns were eased across Europe, more variably among countries than they were imposed (widening confidence intervals in Figure 4b), the decline in deaths stalled. The CHI fell to 50% on average by 15 July by which time the daily growth rate of deaths was once again approaching zero (Figure 4b). From June onwards, the daily growth rate was often above zero, consistent with reports of renewed outbreaks of COVID-19 across the European region, and reinforcing the view that the previous rise in deaths had been constrained, at least in part, by lockdown (Figure 4a).

In conclusion, this investigation differs from others in using an empirical model (skew-logistic, devised for this study), to explore the factors that affect the size of COVID-19 epidemics (measured in terms of reported deaths) across Europe. We find that countries reporting fewer deaths did not generally have lower rates of transmission and epidemic growth; that is, they did not have flatter epidemic curves, driven by lower values of *R_0_* (contrary to Figure 1a). Rather, countries with fewer deaths were those that locked down earlier (having reduced *R_0_* to *R_0c_*) generating shorter epidemics that peaked sooner (Figure 1b). Similar effects of lockdown have been inferred from previous analyses of COVID-19 in European (*3, 13*) and other countries, including China (*16-20*).

Fewer deaths were also reported by countries with fewer inhabitants, for reasons that are not yet clear. If lockdown and non-pharmaceutical interventions were the sole mechanism of epidemic control, we would expect no association between the number of deaths and population size. One possible explanation is that smaller countries had fewer introduced infections from which national epidemics grew. Another is that only a subset of any national population is exposed or susceptible to infection and illness (*9, 10*), and that sub-population is smaller in countries with fewer inhabitants. The distribution of coronavirus is highly heterogeneous in every country, as any map of COVID-19 sub-national distribution shows (*21*). This latter hypothesis implies that epidemics have been controlled, in part, by the local (sub-national) depletion of susceptibles and the build-up of herd immunity (changes in *R_t_* falling below *R_0_*; Figure 1b).

This analysis, like previous studies (*3, 13, 22*), exposes the perils of delayed action during an epidemic with an average doubling time as short as three days. At this rate of growth, the daily death toll would have increased by a factor of ten within nine days; this is about the same as the average time delay from one death to the time of lockdown, which was 9.0 (95%CI 2.6) days. In effect, a COVID-19 epidemic in a typical European country would have expanded more than ten-fold within the average time it took to impose lockdown.

The need for speed is expected to apply to resurgent epidemics too. The strong link between COVID-19 deaths and the time of lockdown implies that only a small fraction of Europe’s population has so far been exposed to infection, a view reinforced by serological surveys that generally report less than a 10% national prevalence of antibody to SARS Cov-2 (*13, 23-25*). Consequently, like other investigators (*1, 3, 11, 14, 26*), we expect that further COVID-19 outbreaks will continue to threaten large numbers of susceptible people across Europe. The apparent effectiveness of lockdowns (Figure 4a), and the penalty for releasing them (Figure 4b), underline the central dilemma facing European countries now: how to maintain the beneficial effects of physical distancing without reinstating full lockdowns across the continent.

As a comparative analysis of European countries — which has identified a strong association between COVID-19 deaths, the timing of lockdown and population size — this study says nothing about the importance of other factors that could affect COVID-19 epidemiology. Among these factors are environmental risks such as air pollution (*27*), underlying health conditions (*28*) including obesity (*29*), possible protective cross-immunity to other coronaviruses (*30, 31*), ethnicity and occupation (*32, 33*), demography and age structure (*34, 35*), and methods of treatment and clinical care that could lower case fatality (*36, 37*). Whether any of these factors can help to explain the epidemiological differences between countries, with a view to finding better ways of containing COVID-19 across Europe, remains an open question.

## Data Availability

All data are publicly available.

## Acknowledgments

CD was supported by a Visiting Fellowship at the Oxford Martin School. We thank Charles Godfray, Moritz Kraemer and Oliver Pybus for helpful suggestions.

## Author contributions

The authors jointly formulated the research goals and aims, designed the study and compiled the data. RCHC, JSD and BGW developed the mathematical and statistical models and estimation routines. CD wrote the first draft of the paper, which was finalized with contributions from all authors.

## Data and materials availability

All data are available in the main text, in the Supplementary Materials, and in the sources cited. Software to fit the skew-logistic model is available from the authors.

## Supplementary Materials

### Materials and Methods

This supplement to the main text provides data sources, methods of data analysis, the mathematical model of COVID-19 dynamics, plus additional data and Figures.

### Data sources

This analysis is based on COVID-19 deaths reported from 30 European countries (*1*) for which the ISO 3166-1 alpha-3 abbreviations are in Table S1. Like other investigators (*2*), we used reported deaths because they are likely to be more accurate than reported cases, both in absolute magnitude (scale of the epidemic) and in the distribution of deaths through time (shape or dynamics of the epidemic). The information on daily deaths is compiled by the European Centre for Disease Prevention and Control using standard methods applied to all countries (*1*).

Although the completeness of death reports has been questioned, the clear patterns of association in Figs 3 and 4 of the main text suggest that both the data, and the empirical model (skew-logistic) used to describe the data, portray real effects, rather than artefacts, of COVID-19 epidemics across the 30 countries included in this investigation.

**Table S1.** Three-letter ISO codes for the 30 countries included in this analysis (Figs 4a, S5, S6).

|  |  |  |  |
| --- | --- | --- | --- |
| Albania | ALB | Italy | ITA |
| Austria | AUT | Luxembourg | LUX |
| Belgium | BEL | Netherlands | NLD |
| Bulgaria | BGR | Norway | NOR |
| Croatia | HRV | Poland | POL |
| Czech Republic | CZE | Portugal | PRT |
| Denmark | DNK | Romania | ROU |
| Finland | FIN | Serbia | SRB |
| France | FRA | Slovak Republic | SVK |
| Germany | DEU | Slovenia | SVN |
| Greece | GRC | Spain | ESP |
| Hungary | HUN | Sweden | SWE |
| Iceland | ISL | Switzerland | CHE |
| Ireland | IRL | Turkey | TUR |
| Israel | ISR | United Kingdom | GBR |

### SEIR model of COVID-19 transmission dynamics

Fig 1 of the main text was constructed with a compartmental model of SARS CoV-2 transmission, framed in ordinary differential equations, and representing a homogeneously mixing population divided among those who are susceptible, exposed, infectious and recovered or died (SEIR), as follows:

**Fig S1.**
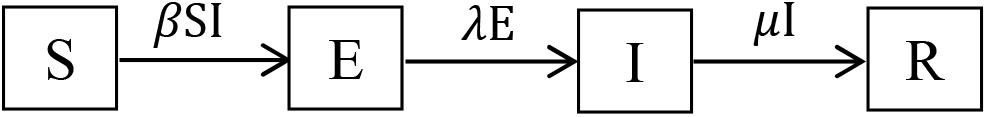
The SEIR model. The compartments denote those in the population that are Susceptible, Exposed, Infected and Recovered.

The variables S, E, I and R satisfy the ordinary differential equations:

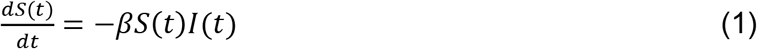

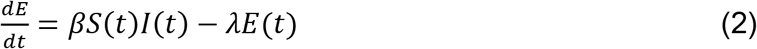

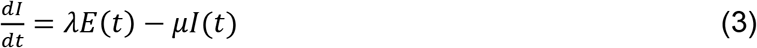

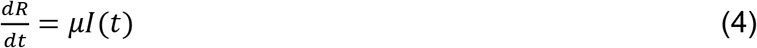

A convenient recent reference is Ma (*3*), though we have adjusted the notation here, reserving certain symbols for use in other standard ways.

**Fig S2.**
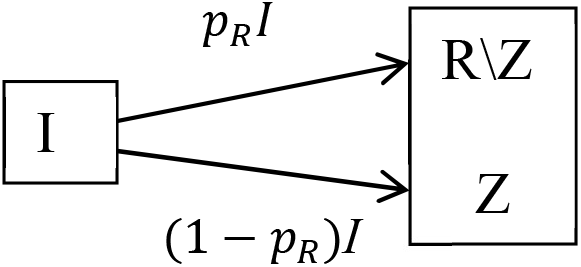
Adjustment of the SEIR model where R is divided into two compartments, R \ Z, those that recover and Z, those that die; where *p_R_* is the proportion that recover.

As this discussion focuses on the number of people that die from COVID-19, the SEIR model is adjusted by splitting R, those that recover, to distinguish between Z, those that die, from those that survive. This arrangement is depicted in Fig S2, and is a simplified version of that used by Dagpunar (*4*) to consider the outcomes of hospitalization.

We allow this adjusted SEIR model to depend on certain parameters with the understanding that once these parameters are known the behaviour of the SEIR model is completely specified. The parameters of the model are described in Table S2, with parameter values estimated for Germany (Fig 1 of main text).

**Table S2.** Parameters of the SEIR model with estimates for Germany.

| Symbol | Definition | Value for Germany in Fig 1 (95%CI) |
| --- | --- | --- |
| $D = \mu^{-1}$ | mean duration of infectiousness | 9.0 (7.9 – 9.9) days<br>or 5.0 or 4.0 (fixed) for comparison (Fig 1a) |
| $T = \beta^{-1}$ | mean interval between infectious contacts | 3.0 (2.8 – 3.3)<br>$R_0 = \beta/\mu = D/T$ |
| $M = \lambda^{-1}$ | mean latent period | 4.0 (2.5 – 5.1) days |
| $t_0$ | number of days from start of epidemic before observations began | 33 days (fixed) |
| $e_0$ | initial number of exposed | 9.2 (7.0 – 10.5) E-05 |
| $s_0$ | initial size of exposed population (to scale the epidemic) | 1.09 (1.07 – 1.15) E+06<br>or 1.5 or 0.5 E+06 for comparison (Fig 1b) |
| $\sigma$ | standard deviation of observational error | 41.9 (34.1 – 44.9) |
| $p_R$ | probability of recovering | 0.991 (fixed) |
| $\omega$ | mean time between the end of infectiousness and death | 11 days (fixed) |

For those who die from COVID-19, parameter *ω* specifies the time from the end of infectiousness to death (Fig S2, equation (4) so that:

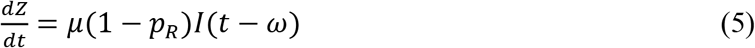

We denote by **θ** = (*b*1*,b*2*,…,b*9), the vector of parameters. With **θ** given, the four differential equations (1), (2), (3) and (5) can be solved by numerical integration to give the trajectories

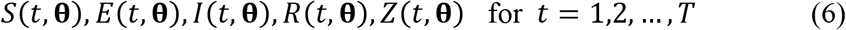

where *t* is the day and *T* is the number of days of interest. We used the standard method of Maximum Likelihood (ML), as given for example in Cheng (*5*), to estimate parameter values.

Here we outline the approach (fitting the model) used to estimate the parameters from a sample of observed daily deaths, this being used to prepare Fig 1 in the main text. Let the sample of observed number of daily deaths be denoted by

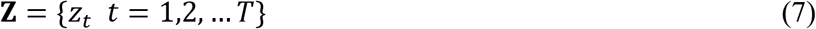

where *z_t_* is the number of deaths on day *t* and *T* is the number of days observed. If the observations were made without error and if, with the right parameter values are correct for **θ**, then the death trajectory {*Z*(*t*, **θ**) *t* = 1,2,…, *T*} would match the observed deaths **Z** in (*7*). So the model would then be successful in explaining deaths.

To include statistical uncertainty in the model we assume instead

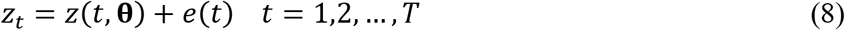

where *e*(*t*) is random error. For simplicity the *e*(*t*) are assumed to be normally and independently distributed (NID) with standard deviation *σ*, i.e.

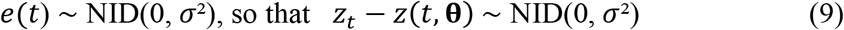

The logarithm of the distribution of the sample is then

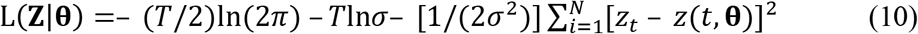

where **Z** is the random argument, and the parameters **θ** are fixed. In ML estimation (MLE), this is turned on its head so that **Z** is simply the known sample of observations now regarded as fixed and we write L as L(**Z|θ**) = L(**θ|Z**)) calling it the (log)likelihood to indicate that it is now treated as a function of **θ**. The ML estimator **θ** is simply the value of **θ** at which L(**θ**|**Z**) is maximized. i.e.

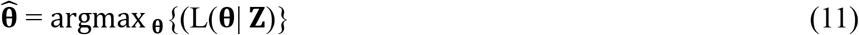

Nelder-Mead numerical search for the maximum was used. This goes through different **θ***_i_*; *i*=1, 2, 3,… comparing the different L(**θ***_i_*,|**Z**) to find 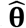, the best **θ**.

To simplify description of the estimation process, only fitting to deaths data, **Z** as in (*7*) has been described, but the method extends straightforwardly to include other data samples. For example

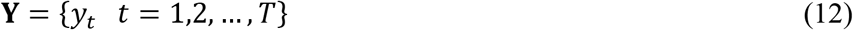

where *y_t_* is the number of active cases on day *t*. Fitting simultaneously to both **Y** and **Z** can be carried out by adding to the right-hand side of (10) a corresponding set of terms for **Y**. Numerical solution of the differential equations requires initial values for *S, E, I, R*. These are essentially scale invariant with (*S + E + I + R*) constant and independent of t. So the numerical integration can conveniently be done using S(0, **θ**) = 1, E(0, **θ**) some small quantity subsequently adjustable as its initial value *e*_0_ is a parameter; with *I* and *R* also initially zero. The size of the exposed population, parameter *s_0_*, is only needed to provide scaled values *S*, *E*, *I*, *R* at each step for comparison with the data **Y** and **Z**.

### Empirical model of epidemic growth and decline: skew-logistic

The SEIR model above cannot accurately describe European COVID-19 epidemics when constrained by biologically plausible parameter values, notably the slow rates of decline in the asymmetric epidemics. Without changing initial conditions, the number of people infected (and the number that die), and the timing of the epidemic, can be adjusted either by changing *R_0_* (Fig 1a) or by changing size of the exposed or susceptible population (Fig 1b). For example, to describe the epidemic in Germany (Fig 1), the SEIR model above assumes that only a fraction of a national population (*s_0_*) is exposed to or susceptible to infection prior to the introduction of COVID-19. The size of the epidemic is then determined by *R_0_* and by the rate of decline in *R_t_* as the susceptible population is depleted by the spread of infection. The alternative way to describe the European epidemics is to allow a change in initial conditions, in particular by reducing *R_0_* to *R_0c_* during the course of the epidemic, for example to represent the effect of non-pharmaceutical interventions) (*2, 6, 7*).

In order to avoid making strong assumptions *a priori* about the mechanism of epidemic control, we devised a flexible empirical model to describe the European epidemics. A number of functional forms (variations on the skew-logistic, the Shannon entropy index (*8*), and others) could probably describe these data but, for this study, we derived a new form of the skew-logistic:

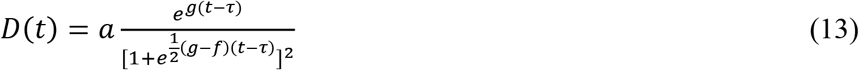

where *D*(*t*) is the number of deaths per unit time (although the model could apply to case incidence too). The model depends on just four parameters: *a*, a scaling parameter used to adjust the size of the epidemic; *g* and *f*, two slope parameters, one characterising the rate of rise, the other the rate of decline of the epidemic; and *τ*, a location time parameter indicating when the epidemic is at its peak.

We assume that *g >* 0 and *f <* 0 so that the term 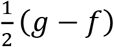 in the exponential expression in the denominator is always positive. Then *D*(*t*) ≅ *e^a^*^(^*^t−τ^*^)^ if *t →* −∞, so that g gives the exponential rate of increase of the epidemic as t increases from —ro. However, *D*(*t*) ≅ *e^f^*^(^*^t−τ^*^)^ if *t → ∞*, sc that *f <* 0 gives the exponential rate of decrease of the epidemic as this subsides.

In addition, equation (13) is identical to

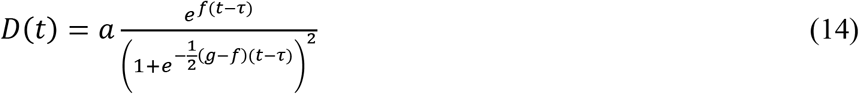

So that *g* and *f* are mathematically identical, and either (13) or (14) can be used when fitting to data, with *g* and *f* playing either role depending on their sign.

There are two useful additional characteristics of *D*(*t*). First, it has the explicit maximum value

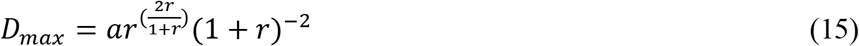

where *r = −g/f* Second, the maximum point is at

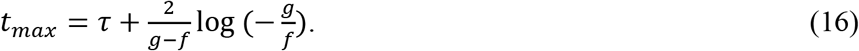

This shows that *τ* is close to where the epidemic maximum occurs, if *g* and −*f* are comparable so that the second term in the right-hand side of (16) is near zero. In any case, *t_max_* indicates where the epidemic will peak; a convenient quantity for comparing the epidemics in different countries. In addition, we can measure the period of epidemic growth as the time from 1 death/day (estimated from the model fit rather than counted in the data) to *t_max_*.

Thus all four parameters have readily understandable, practical interpretations, and the model can easily be fitted to data (cases or deaths) using the method of maximum likelihood (ML), where we employ here Newton-Raphson numerical search. This latter needs initial parameter values to be provided. We do this by first examining the data to see if the epidemic appears to have peaked or not, and if so obtain an initial estimate of its position and size. This allows rates of increase and decrease to be roughly estimated to give starting values for *g* and *f*. An initial value for *a* can then be obtained using (15), whilst equation (16) shows that the observed maximum of the epidemic can be used as the initial value of *τ*. Note that an extra parameter, *σ*, the standard deviation of the observational error, is included in the ML method, and this parameter also being estimated.

This skew-logistic model should not be mistaken for the established, similarly named, skew-logistic distribution and its generalizations, these latter being probability distributions that have different mathematical characteristics to (13). The new model is particularly suited to bell-shaped but asymmetric epidemic trends where there is a rapid increase to a peak before a decrease that is usually more gradual.

We fitted the model to data from 24 countries for which there were sufficient reported deaths (Figs 2 and 3, Fig S3), giving the ML estimates in Fig 2 and Table S3 (at the end of this document). Software to fit the model is available from the authors.

**Fig S3.**
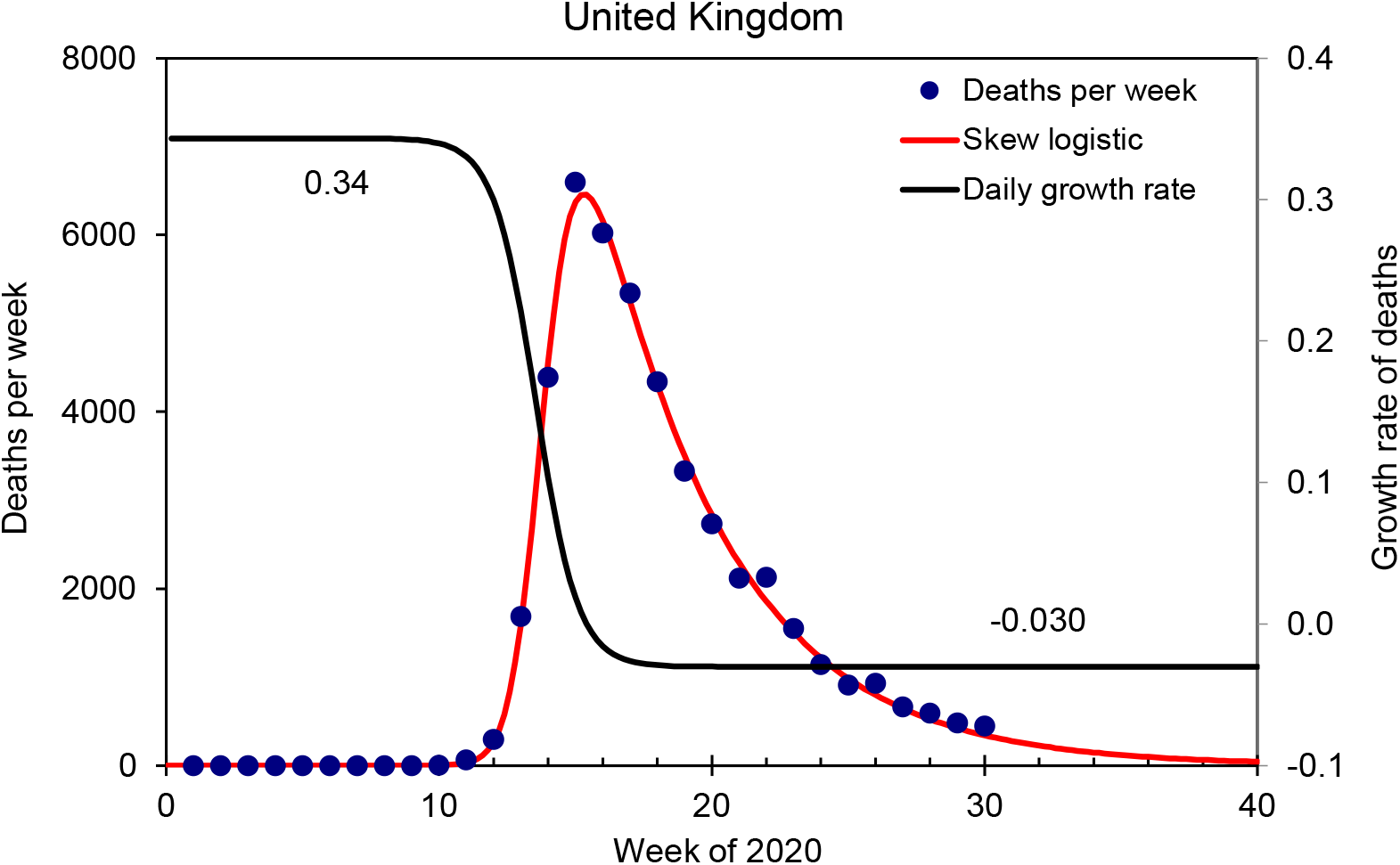
Estimates of the rates of epidemic growth and decline obtained from the skew-logistic model, illustrated for the United Kingdom. Data (points) are summed over 7 days, covering the weekly reporting cycle. The maximum growth rate of the epidemic is 0.34/day (the growth rate that determines *R_0_*) and the maximum rate of decline is 0.03/day. The fit is also shown in Fig 2 of the main text.

### Non-pharmaceutical interventions: policies on lockdown

We characterized lockdown policies based on the Government Response Tracker created by the Blavatnik School of Government, Oxford (*9*). The Containment and Health Index is based on eleven indicators including containment and closure policies (schools, workplaces, travel bans) and health system policies (information, testing, contact tracing), with a score for each country varying between 0 and 100%.

For each country, CHI increased with time (Fig S4) and, in general, later decreased (Fig 4b). Using this index, we calculated the cumulative number of deaths at lockdown (286 deaths in the UK) as the average number weighted by the magnitude of incremental step of CHI. The date of lockdown is the date corresponding to the average of the cumulative number of deaths (22-23 March in the UK).

**Fig S4.**
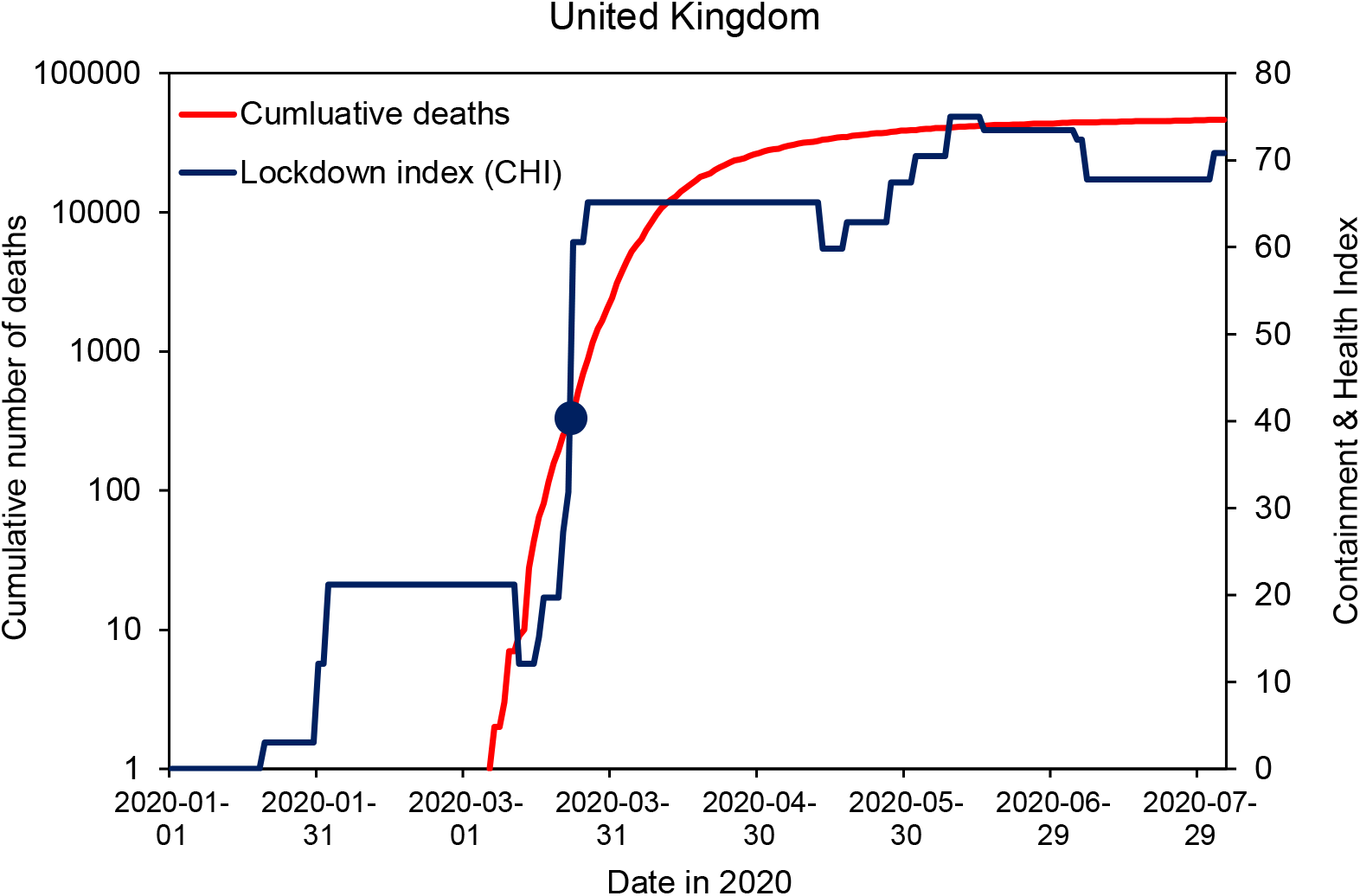
Cumulative reported deaths (red) and the Containment and Health Index (blue) for the United Kingdom. The circle marks the number of deaths (286) and the corresponding date of lockdown (22-23 March).

**Fig. S5.**
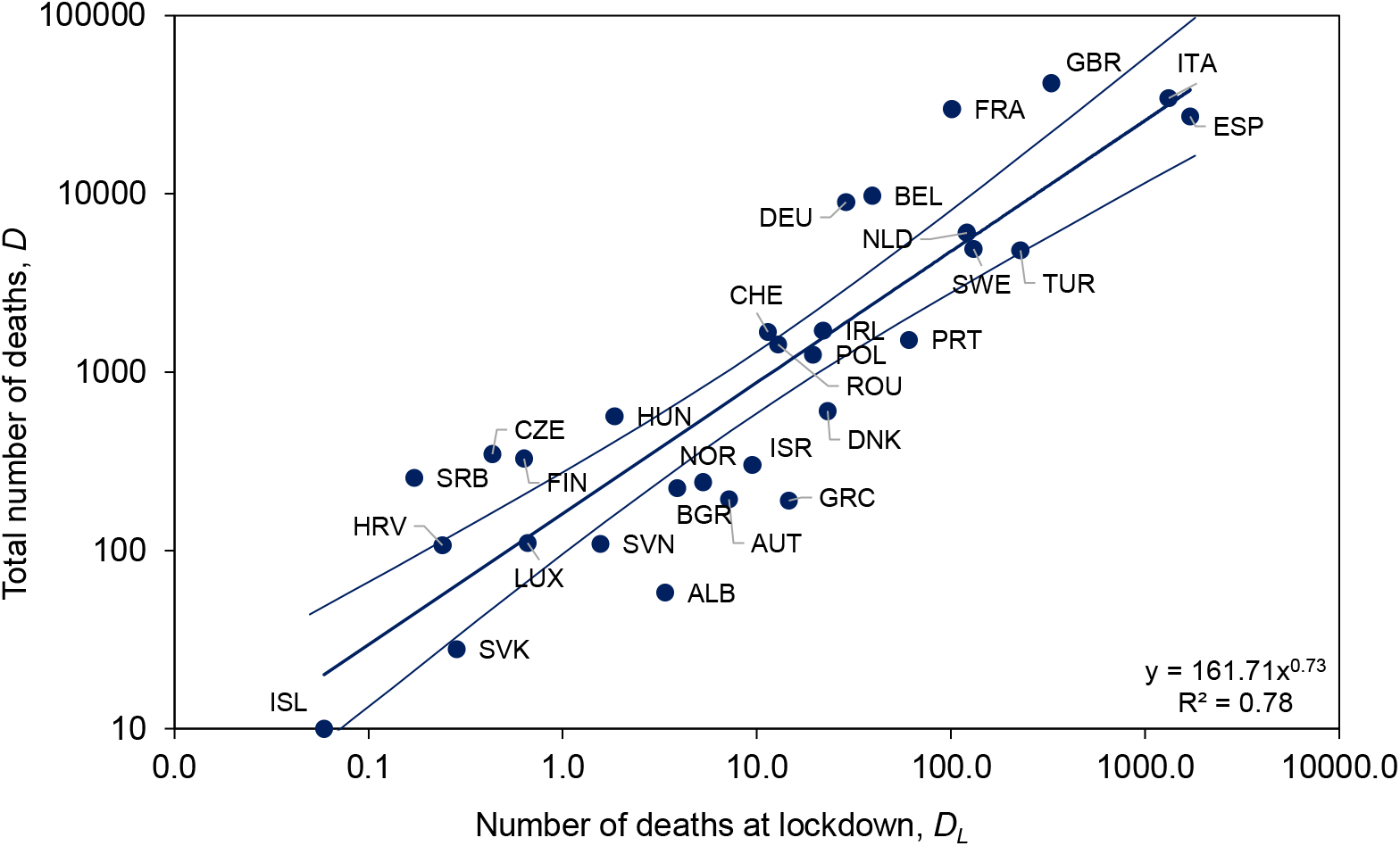
The relation between total deaths and deaths at lockdown based on CHI, as in Fig 4a of the main text. The power relationship (central line) is 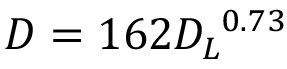. Errors on the regression line are 95%CI (lateral lines).

This interpretation of the number of deaths at lockdown gives the univariate relation between total deaths (*D*) and deaths at lockdown (*D_L_*) shown in Fig S5, which is integral to the multiple regression in Fig 4a of the main text.

Alternative interpretations of the date of lockdown give similar results. One of these alternatives, among several permutations explored, is the number of deaths recorded when the Stringency Index (as for CHI, but excluding the health system policies) reached 70% (Fig S6) (*9*).

**Fig. S6.**
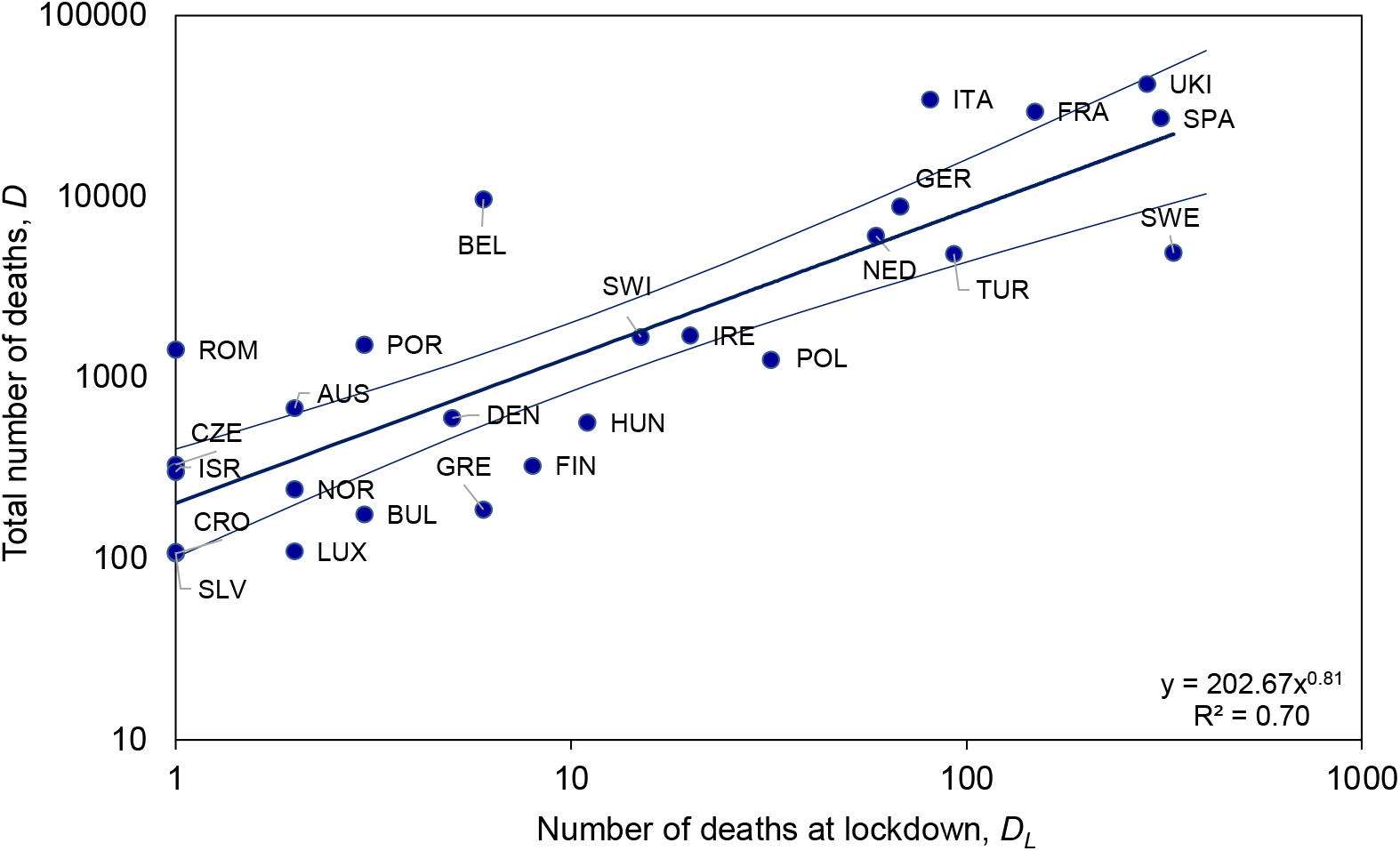
An alternative interpretation of the timing of lockdown based on the Stringency Index applied to the 24 countries in Figs 2 and 3, yielding similar results to Fig S5. The power relationship (central line) is 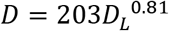. Errors on the regression line are 95%CI (lateral lines).

**Table S3.** Parameter estimates for the skew-logistic model fitted to 24 countries.

| Time units are days; confidence Intervals are 95% |  |  |  |  |  |  |  |  |  |  |  |  |  |  |  |  |  |  |  |  |  |  |  |  |  |
| --- | --- | --- | --- | --- | --- | --- | --- | --- | --- | --- | --- | --- | --- | --- | --- | --- | --- | --- | --- | --- | --- | --- | --- | --- | --- |
| Country | σ |  |  |  |  | a |  |  |  |  | g |  |  |  |  | f |  |  |  |  | τ |  |  |  |  |
|  | MLE | Asymp Lo | Asymp Hi | BS Lo | BS Hi | MLE | Asymp Lo | Asymp Hi | BS Lo | BS Hi | MLE | Asymp Lo | Asymp Hi | BS Lo | BS Hi | MLE | Asymp Lo | Asymp Hi | BS Lo | BS Hi | MLE | Asymp Lo | Asymp Hi | BS Lo | BS Hi |
| Austria | 1.1 | 0.8 | 1.3 | 1.0 | 1.2 | 56.1 | 42.1 | 70.1 | 47.8 | 65.3 | 0.23 | 0.16 | 0.30 | 0.20 | 0.26 | -0.06 | -0.08 | -0.05 | -0.08 | -0.05 | 1.92 | 1.84 | 2.00 | 1.88 | 1.98 |
| Belgium | 4.2 | 3.1 | 5.2 | 3.6 | 4.8 | 821.3 | 770.6 | 872.1 | 791.2 | 851.1 | 0.25 | 0.23 | 0.27 | 0.24 | 0.26 | -0.07 | -0.07 | -0.06 | -0.07 | -0.06 | 1.95 | 1.94 | 1.97 | 1.94 | 1.96 |
| Croatia | 0.3 | 0.2 | 0.4 | 0.3 | 0.4 | 10.4 | 7.9 | 12.8 | 7.9 | 12.5 | 0.11 | 0.04 | 0.18 | 0.08 | 0.14 | -0.09 | -0.14 | -0.03 | -0.14 | -0.06 | 2.34 | 2.00 | 2.69 | 2.19 | 2.51 |
| Czechia | 0.5 | 0.4 | 0.7 | 0.5 | 0.6 | 16.2 | 13.2 | 19.1 | 13.5 | 19.1 | 0.43 | 0.29 | 0.56 | 0.36 | 0.51 | -0.04 | -0.05 | -0.03 | -0.05 | -0.03 | 1.88 | 1.85 | 1.91 | 1.86 | 1.91 |
| Denmark | 0.6 | 0.4 | 0.7 | 0.5 | 0.7 | 31.0 | 26.8 | 35.2 | 28.0 | 34.2 | 0.33 | 0.25 | 0.41 | 0.29 | 0.37 | -0.04 | -0.05 | -0.04 | -0.05 | -0.04 | 1.84 | 1.82 | 1.87 | 1.83 | 1.86 |
| Finland | 0.9 | 0.7 | 1.2 | 0.8 | 1.1 | 39.6 | 28.8 | 50.3 | 31.9 | 44.7 | 0.18 | 0.08 | 0.29 | 0.14 | 0.29 | -0.11 | -0.16 | -0.06 | -0.19 | -0.08 | 2.36 | 2.21 | 2.51 | 2.29 | 2.46 |
| France | 22.6 | 16.9 | 28.4 | 20.0 | 25.4 | 2178.6 | 1934.6 | 2422.6 | 2033.5 | 2342.9 | 0.34 | 0.28 | 0.39 | 0.30 | 0.37 | -0.06 | -0.07 | -0.05 | -0.07 | -0.06 | 1.92 | 1.90 | 1.94 | 1.91 | 1.93 |
| Germany | 4.6 | 3.4 | 5.7 | 3.8 | 5.4 | 635.3 | 583.9 | 686.7 | 601.6 | 669.7 | 0.20 | 0.18 | 0.22 | 0.19 | 0.21 | -0.06 | -0.06 | -0.05 | -0.06 | -0.05 | 2.03 | 2.01 | 2.06 | 2.02 | 2.05 |
| Greece | 0.4 | 0.3 | 0.5 | 0.3 | 0.4 | 7.3 | 5.6 | 9.0 | 5.7 | 9.0 | 0.41 | 0.22 | 0.61 | 0.35 | 0.51 | -0.03 | -0.04 | -0.03 | -0.04 | -0.03 | 1.71 | 1.67 | 1.75 | 1.68 | 1.74 |
| Hungary | 0.4 | 0.3 | 0.5 | 0.4 | 0.5 | 26.3 | 23.3 | 29.3 | 23.7 | 29.5 | 0.29 | 0.23 | 0.34 | 0.25 | 0.33 | -0.04 | -0.04 | -0.03 | -0.04 | -0.03 | 2.05 | 2.02 | 2.07 | 2.03 | 2.07 |
| Israel | 0.5 | 0.4 | 0.7 | 0.5 | 0.6 | 17.9 | 13.7 | 22.2 | 15.0 | 21.8 | 0.34 | 0.22 | 0.45 | 0.29 | 0.39 | -0.05 | -0.06 | -0.04 | -0.06 | -0.04 | 1.95 | 1.91 | 1.99 | 1.92 | 1.98 |
| Italy | 16.2 | 12.1 | 20.3 | 13.3 | 19.9 | 1554.6 | 1450.0 | 1659.2 | 1452.1 | 1648.7 | 0.29 | 0.26 | 0.32 | 0.27 | 0.31 | -0.04 | -0.04 | -0.03 | -0.04 | -0.03 | 1.64 | 1.62 | 1.65 | 1.62 | 1.65 |
| Luxembourg | 0.3 | 0.2 | 0.4 | 0.2 | 0.3 | 8.0 | 5.3 | 10.7 | 6.3 | 9.8 | 0.25 | 0.14 | 0.36 | 0.21 | 0.30 | -0.06 | -0.08 | -0.04 | -0.07 | -0.05 | 1.85 | 1.76 | 1.94 | 1.80 | 1.90 |
| Netherlands | 5.0 | 3.7 | 6.2 | 4.1 | 6.1 | 395.2 | 341.9 | 448.5 | 350.6 | 443.0 | 0.22 | 0.18 | 0.27 | 0.19 | 0.26 | -0.05 | -0.06 | -0.04 | -0.06 | -0.05 | 1.89 | 1.85 | 1.93 | 1.86 | 1.93 |
| Norway | 0.4 | 0.3 | 0.5 | 0.3 | 0.4 | 22.4 | 17.2 | 27.5 | 19.7 | 25.4 | 0.22 | 0.15 | 0.29 | 0.19 | 0.24 | -0.07 | -0.09 | -0.05 | -0.09 | -0.06 | 2.02 | 1.94 | 2.10 | 1.99 | 2.06 |
| Poland | 1.9 | 1.4 | 2.4 | 1.6 | 2.5 | 27.8 | 23.4 | 32.3 | 21.7 | 36.2 | 0.38 | 0.20 | 0.56 | 0.29 | 0.52 | -0.01 | -0.01 | -0.01 | -0.02 | -0.01 | 1.94 | 1.89 | 1.99 | 1.91 | 1.98 |
| Portugal | 2.3 | 1.7 | 2.9 | 2.1 | 2.8 | 52.5 | 42.1 | 62.8 | 40.9 | 70.8 | 0.31 | 0.20 | 0.42 | 0.25 | 0.40 | -0.03 | -0.03 | -0.02 | -0.03 | -0.02 | 1.84 | 1.79 | 1.88 | 1.80 | 1.90 |
| Romania | 2.2 | 1.6 | 2.8 | 1.7 | 2.8 | 40.6 | 27.1 | 54.1 | 25.7 | 65.5 | 0.24 | 0.11 | 0.37 | 0.15 | 0.49 | -0.02 | -0.02 | -0.01 | -0.03 | -0.01 | 1.93 | 1.85 | 2.01 | 1.87 | 2.07 |
| Slovenia | 0.2 | 0.2 | 0.3 | 0.2 | 0.3 | 8.7 | 6.5 | 11.0 | 7.2 | 10.4 | 0.29 | 0.19 | 0.39 | 0.25 | 0.34 | -0.06 | -0.08 | -0.05 | -0.08 | -0.05 | 1.94 | 1.88 | 1.99 | 1.90 | 1.97 |
| Spain | 27.6 | 20.0 | 35.3 | 22.7 | 33.1 | 1791.0 | 1564.8 | 2017.1 | 1599.9 | 1955.6 | 0.35 | 0.28 | 0.42 | 0.31 | 0.40 | -0.05 | -0.06 | -0.04 | -0.05 | -0.05 | 1.77 | 1.75 | 1.79 | 1.75 | 1.79 |
| Sweden | 4.3 | 3.2 | 5.4 | 3.8 | 5.3 | 165.2 | 144.9 | 185.5 | 140.6 | 192.9 | 0.24 | 0.19 | 0.29 | 0.21 | 0.27 | -0.02 | -0.03 | -0.02 | -0.03 | -0.02 | 1.99 | 1.96 | 2.03 | 1.96 | 2.03 |
| Switzerland | 2.1 | 1.5 | 2.6 | 1.7 | 2.5 | 126.5 | 100.2 | 152.7 | 107.3 | 148.1 | 0.22 | 0.15 | 0.28 | 0.17 | 0.27 | -0.06 | -0.07 | -0.05 | -0.07 | -0.05 | 1.89 | 1.82 | 1.96 | 1.84 | 1.96 |
| Turkey | 3.8 | 2.7 | 4.9 | 3.2 | 4.5 | 286.3 | 247.9 | 324.7 | 259.4 | 316.4 | 0.23 | 0.19 | 0.27 | 0.21 | 0.25 | -0.05 | -0.05 | -0.04 | -0.05 | -0.04 | 2.04 | 2.00 | 2.07 | 2.01 | 2.07 |
| United Kingdom | 14.6 | 10.9 | 18.4 | 13.0 | 17.2 | 1615.8 | 1544.9 | 1686.7 | 1539.3 | 1693.8 | 0.34 | 0.32 | 0.37 | 0.33 | 0.36 | -0.03 | -0.03 | -0.03 | -0.03 | -0.03 | 1.92 | 1.91 | 1.93 | 1.91 | 1.93 |

The skew-logistic model is fitted to data on the number of deaths reported daily as these should have more independent and identically distributed random errors than 7-day moving averages (Table S3). Upper and lower confidence limits were obtained both by standard asymptotic theory and bootstrapping. Asymptotic theory tends to give more optimistic (narrower) confidence intervals (CI). Bootstrapping allows for asymmetry in the CI, which are wider.

The countries listed in this Table exclude Albania, Bulgaria, Iceland, Ireland, Serbia and Slovakia for which epidemics could not be described by the skew-logistic, either because there were too few data (e.g. Iceland), or the data did not describe a standard epidemic curve (Bulgaria).

